# An intersectional study of poverty, migration, and treatment seeking behaviour on antimicrobial use in a semi-urban area in Tamil Nadu, India

**DOI:** 10.64898/2026.04.29.26352106

**Authors:** Vijayaprasad Gopichandran, Neha Muralidharan, Jayasree Chandrasekaran, Darcus Deva Sinthiya, Sudharshini Subramaniam, Rajeswaran Thiagesan, Jolly Ranjith

**Affiliations:** Independent Community Health Consultant, Chennai, Tamil Nadu, India; Omayal Achi Community Health Centre, Arakkampakkam, Thiruvallur, Tamil Nadu, India; Department of Community Health Nursing, Omayal Achi College of Nursing, Puzhal, Chennai, Tamil Nadu, India; Department of Community Medicine, Government Vellore Medical College, Vellore, Tamil Nadu, India; Resource Group for Education and Advocacy in Community Health, Chennai, Tamil Nadu, India; Department of Medical and Surgical Nursing, Omayal Achi College of Nursing, Puzhal, Chennai, Tamil Nadu, India

**Keywords:** antimicrobial use, intersectionality, socioeconomic status, migration, private healthc are, antimicrobial stewardship

## Abstract

**Background:** Understanding antimicrobial use and factors driving it in communities is essential to devise its stewardship and reduce emergence of antimicrobial resistance.

**Objectives:** To study the intersectional influence of socioeconomic status, migration and place of treatment on antimicrobial use in a semi urban area in Tamil Nadu, India.

**Methods:** We conducted a cross-sectional survey among systematically sampled 525 adult men and women from three villages in a semi-urban area in Tiruvallur district. We collected data through structured interviews on incidence of infections in the past 3 months, treatment seeking behaviour, and audited the antimicrobial prescription or empty packs of medicines used. We analyzed the data using R statistical software and performed a multilevel analysis of individual heterogeneity and discriminatory accuracy to study intersectional effects.

**Results:** We found that the incidence of infection syndrome was 37% with a majority of them being acute respiratory infections. 143 of them sought treatment, with 40% going to a private general practitioner. People belonging to middle class had a 3.7 times greater odds of going to private sector compared to lower class. Twenty eight (19.6%) of those who sought treatment received an antimicrobial prescription. Sixty percent of them belonged to Access group, 35.7% Watch and 3.6% Restrict group. There was a significant intersectional effect showing middle class- non migrant – private care seekers having 22% probability of antimicrobial use versus lower class – migrant – government care seekers having 16% probability. The variance partition coefficient was 2.6% showing a small by significant portion of the variance contributed by intersectional identities.

**Conclusion:** Antimicrobial use in the community is significantly shaped by the intersection of socioeconomic status, migrant status and place of seeking care for the infection. Regulation of private sector prescription patterns and improving access to health care for migrants are key policy interventions.

## Introduction

Antimicrobial resistance (AMR) has emerged as a major global health threat, compromising the effectiveness of drugs used to treat infectious diseases. In 2019 alone, 1.27 million deaths were directly attributable to AMR, with 4.95 million deaths associated with drug-resistant infections worldwide. The burden is particularly high in low- and middle-income regions such as sub-Saharan Africa and South Asia, driven by high infection rates and limited healthcare resources. If left unchecked, AMR could cause up to 10 million deaths annually by 2050, surpassing many major causes of mortality.[1]

Inappropriate antimicrobial use, including self-medication, overuse, underuse, and incomplete treatment, drives resistance by promoting the emergence and spread of resistant pathogens. Without intervention, this leads to rising resistance, higher healthcare costs, and worse health outcomes.[2] A substantial proportion of antibiotic use in low- and middle-income countries occurs at the community level, often outside formal health systems. Across study sites, 35% of antibiotics were obtained without a prescription, and 37% of households reported self-medication. Easy access to antibiotics, particularly through retail pharmacies serving as first points of care, combined with convenience, cost, and time constraints, drives high levels of inappropriate use.[3]

In India, high infection burden and widespread antibiotic use create ideal conditions for AMR. Multidrug-resistant infections are prevalent, while antibiotics are among the most consumed globally, shaped by over-the-counter access, self-medication, and variability in prescribing behaviors.[4] Structural factors, including reliance on informal healthcare providers and weak regulatory enforcement of prescription-only rules, further exacerbate inappropriate antibiotic use.[5]

The phenomenon of inappropriate antimicrobial use is influenced by several social structures such as socio-economic status, gender, caste, and migration. There is significant evidence that gender plays a crucial role in influencing antimicrobial use and emergence of antimicrobial resistance. Gendered household roles such as managing water, sanitation, and caregiving further increase exposure to infections and potential drug-resistant pathogens.[6–9] Vulnerabilities also arise during menstruation and unclean childbirth practices.[10–13] Incidence of invasive infections are higher among those who live in poverty as it influences access to safe drinking water, nutritious food, sanitation facilities and leads to overcrowded living conditions. Poverty deprives people of access to good quality health care. This leads to delayed diagnosis, ineffective treatment, inappropriate use of antimicrobials all of which can contribute to antimicrobial resistance.[14] Migration is another major determinant of health and disease. Socio-economic vulnerability, language and cultural barriers and challenges in social acceptance can all adversely affect access to effective health care. This can lead to high burden of infections, inadequate access to health care and inappropriate antimicrobial use.[15,16]

Most evidence examines these social determinants in isolation, but intersectionality highlights how multiple overlapping social identities such as gender, caste, class, age, and migration status interact within structures of power to shape health outcomes. [17] Examining a single variable is insufficient because health outcomes and AMR are influenced by intersecting social determinants, including socioeconomic status, cultural norms, and geographic location. [18]

Current AMR interventions often fail because they focus on facility-based surveillance and hospital-centric strategies, overlooking structural and social determinants such as poverty, gendered power relations, inequitable WASH access, and household decision-making.[19] Effective interventions require community-level, context-sensitive evidence that integrates local norms, lived experiences, and multi-sectoral strategies.[20] Understanding the intersectional influence of poverty, migration, caste, and gender is essential to design interventions that are appropriate, acceptable, and sustainable. This study was conducted to explore the intersectional influence of social determinants such as, socioeconomic status, migration and place of treatment on antimicrobial use in a semi urban area in Tamil Nadu, India.

## Methods

### Study design and setting

This cross sectional study was conducted in three villages in the Villivakkam block of Thiruvallur district in Tamil Nadu India. Thiruvallur district is a rapidly industrializing district in Tamil Nadu. The Ambattur, Avadi and Villivakkam areas in this district share a border with the adjoining capital city of Chennai and and some parts of this area are subsumed under the Greater Chennai Corporation. This area is an industrial hub with many manufacturing units. The area where this study was conducted is close to the Avadi Heavy Vehicle Factory which manufacture military grade heavy vehicles for the Indian Army. Tamil Nadu has some of the best health indicators in the country and has a robust public health system as well as a thriving private health sector.

### Study participants

Based on a systematic review and meta analysis of antimicrobial prescription practices at the primary care level in India, the prevalence was found to be 65%.[21] Using this prevalence data for a 95% confidence level and 10% relative precision, the sample size was calculated as 215. To account for the purposive sampling of the villages we used a design effect of 2, increasing the sample size to 430. Accounting for a 25% non response rate we inflated this to 508 and rounded it off to 510.

Adult men and women living in the three villages were eligible to participate in the study. These three villages are part served by the community health nursing department and the community health centre associated with the nursing college. They were purposively selected as they have a high proportion of migrants working in the local brick kilns and factories. The survey was conducted by the third and fourth year BSc Nursing students as part of their community health posting. The researcher gave intensive training to these students over two days. During this training they introduced the concept of antimicrobial resistance, antimicrobial overuse, social determinants and intersectionality. The students were also trained on the method of obtaining informed consent and administering the survey. The trainers went through each item in the questionnaire and provided standard procedures for administering the item. They also were given an opportunity to practice administering the questionnaire as a pilot test during the training.

The students were taken in small batches of 25 each to the villages. The tutors and senior faculty accompanied the students. They supervised the data collection. Each batch of students was taken to a central place and each student in the batch was allotted 5 households in the village. The supervisor enumerated the total number of houses in the village and appropriately calculated the sampling interval to select the houses to be visited by each student. If a household was locked or there was no response, the immediate next house was allocated to the student. The students obtained written informed consent after explaining the details of the survey to a randomly selected adult in that household. The survey was administered in a pen and paper format. A total of 105 students, 60 from the third year batch and 45 from the final year batch, conducted the survey. The students submitted the filled forms to the supervisor each day after completion of the survey. The supervisor reviewed the forms, identified any incomplete data and alerted the student about this so that they could complete it during the next day of field work.

### Study instrument and variables

We developed a structured questionnaire which comprised of six parts. The first two parts collected details of socio demographic characteristics and variables of the Modified Kuppuswamy Scale for measuring socio-economic status.[22] Part 3 collected details of migration status, inter and within state migration, and reason for migration. Part 4 use a syndromic infectious disease surveillance checklist to collect information regarding the presence of any of the common infectious disease syndromes in the past 3 months.[23] If a person had more than one infection in the past 3 months, the most recent infection was considered for the survey. If they had an infection, they proceeded to part 5 of the questionnaire which sought information on the treatment sought for this infection. If no treatment was sought no further information was collected. If treatment was sought, the place of treatment was inquired. Further they were asked if they received a prescription. If a prescription was present, or if the empty blister packs of medicines were available then it was audited to examine whether an antibiotic was prescribed, as the part 6 of the questionnaire.

The details such as name of antibiotic, dose and duration were also collected from the prescription audit. Where a prescription or empty packs were unavailable, it was considered as antibiotics not used. The survey was administered by the students in Tamil language which was the common language for the locals and intra-state migrants. In case of the inter-state migrants, the survey was administered in Hindi by some of the students and supervisors who were fluent in this language. The survey was conducted during the months of Jan-Feb 2026.

### Bias reduction strategies

Household were selected by systematic random sampling and allotted to the students to prevent selection bias. To prevent recall bias we restricted infections to past 3 months and only details of most recent infection episode were collected. Where there was lack of prescription or empty packs we did not consider them as using an antibiotic.

### Statistical analysis

We calculated the burden of infection in the sample over the past 3 months as incidence percentage with 95% confidence intervals. Among those who had any infection, we computed the treatment seeking behaviour, and distribution of place of seeking treatment. We categorized the place of treatment into government and private health facilities and performed a multivariable logistic regression analysis to study the influence of social determinants such as gender, caste, socioeconomic status and migration on place of treatment. We estimated the incidence of antimicrobial prescription. We performed a heat-map visualization to study the influence of gender, caste, socioeconomic status and migration on antimicrobial prescription. We also analyzed the types of antimicrobials used, and classified them according to the WHO AWaRe classification. We then performed a Multilevel Analysis of Individual Heterogeneity and Discriminatory Accuracy (MAIHDA) to examine the intersectional influence of socioeconomic status, migration and place of treatment on antimicrobial prescription.[24] Antimicrobial prescription was treated as a binary outcome variable. We constructed intersection strata based on socioeconomic status (middle class and lower class), migration status (yes and no) and place of treatment (government and private). We fit multi-level models where the null model calculated the total variance in antimicrobial prescription, fixed effects model which studied the independent effects of socioeconomic status, migration and place of treatment, and a random effects model where the intercept was allowed to vary across the intersectional strata. We calculated and interpreted the Variance Partitioning Coefficient (VPC) to determine the total variance of antimicrobial prescription that was attributable to the variance of the intersectional strata, and predicted probability of antimicrobial prescription in the fixed and random effects models. Data analysis was performed using R statistical software. We used the lme4 package in R to run the MAIHDA model. Missing data of continuous variables were imputed with the mean value, wherever the response of a categorical variable was missing or ‘not mentioned’ we handled it as a missing variable and excluded it from analysis. The code and analysis outputs are available in the GitHub repository https://vijaygopichandran-cmd.github.io/Antimicrobial-data-analysis/

### Ethical considerations

The research proposal was approved after full board review by the institutional ethics committee of International Centre for Collaborative Research, Chennai date 11 July 2025. Written informed consent was obtained from all participants before the survey.

## Results

Of the 550 individuals approached for the survey, 525 agreed to participate giving a response rate of 95.4%. Though our calculated sample size was only 510, we included 525 so that each of the 105 students could meet and survey 5 household members. The characteristics of the participants in the study are shown in Table 1. The sample was predominantly women, with mean age of 46.54 years, belonging to the Hindu religion. Majority of the surveyed participants belonged to scheduled castes, with a 13% with no formal schooling. Most of them were married (81%) and 65% belonged to the lower socioeconomic status. Of the surveyed sample 20% had migrated to the current location from a different district within Tamil Nadu and 1.3% had migrated into the village from another state in India.

**Table 1:**
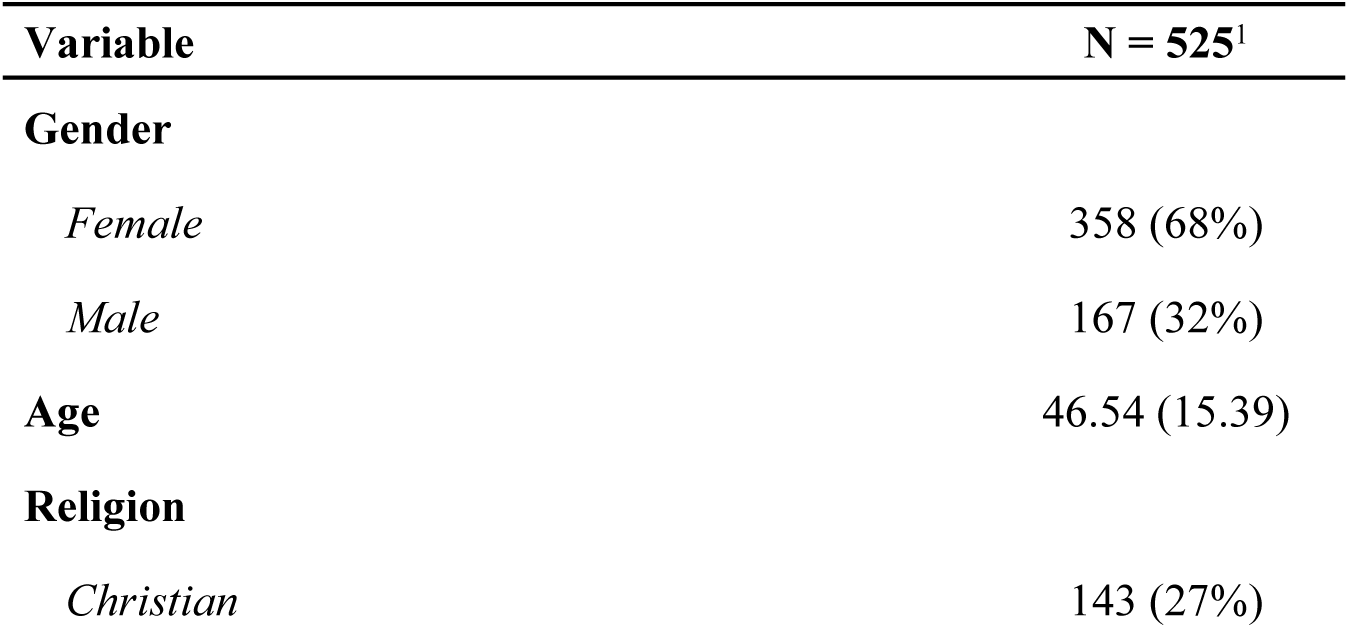
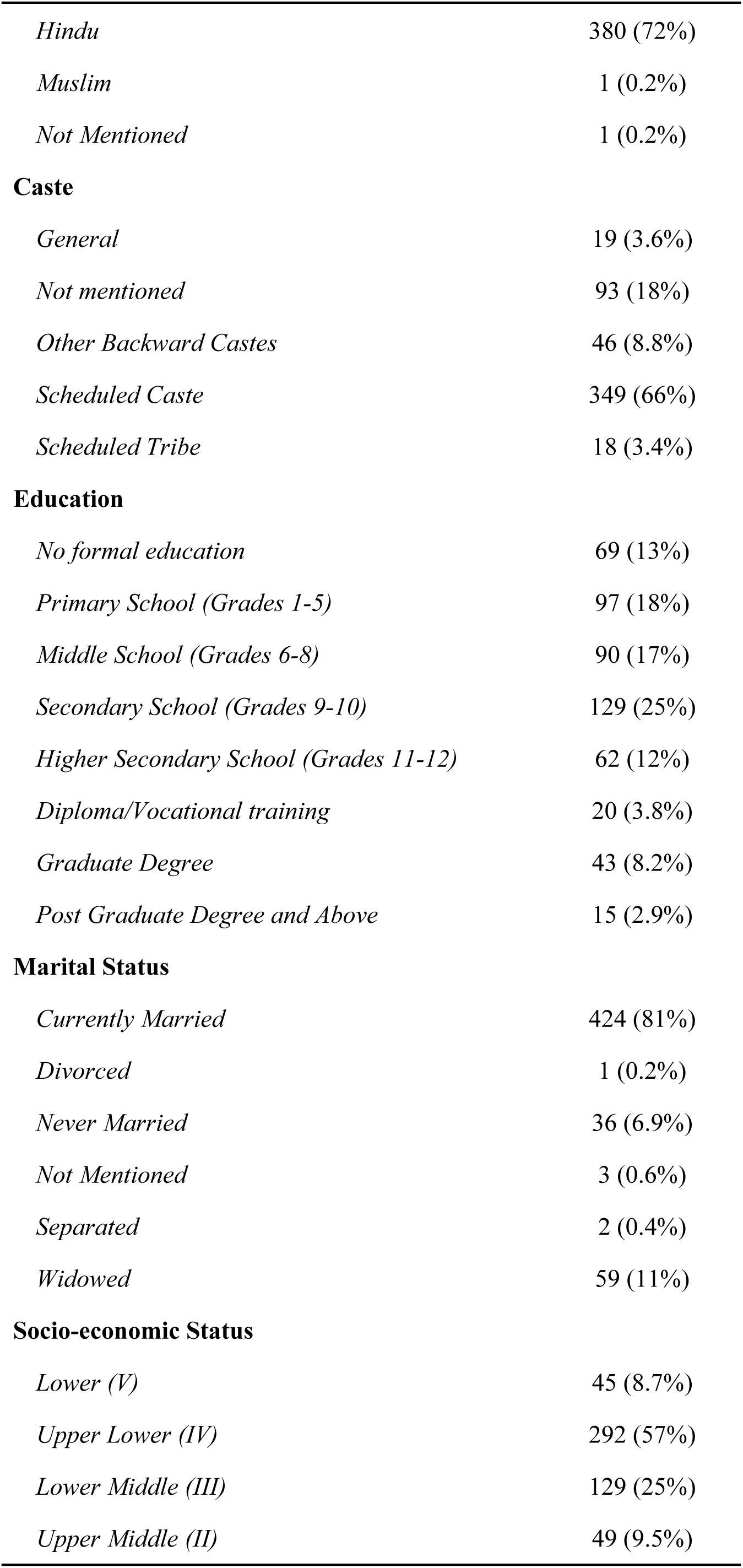
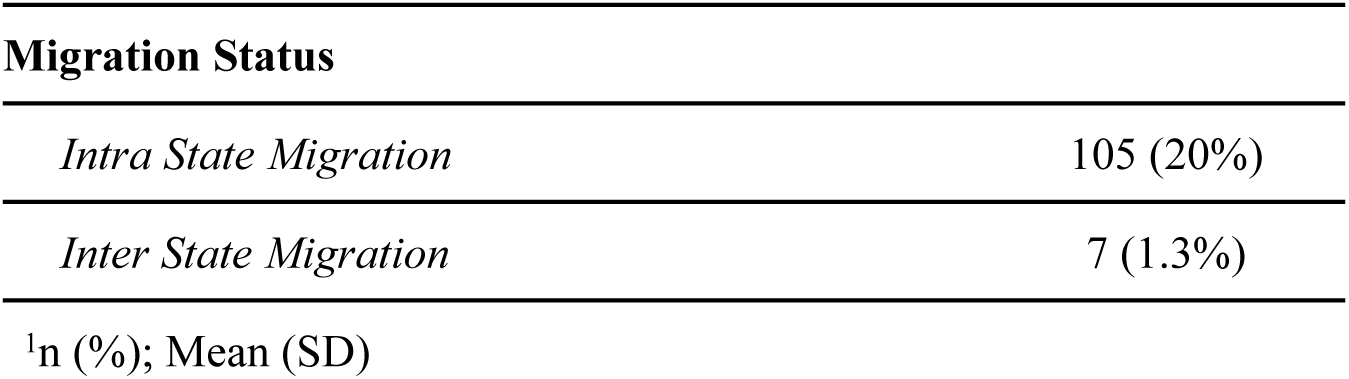
Characteristics of the study sample.

| Variable | N = 525<sup>1</sup> |
| --- | --- |
| **Gender** |  |
| *Female* | 358 (68%) |
| *Male* | 167 (32%) |
| **Age** | 46.54 (15.39) |
| **Religion** |  |
| *Christian* | 143 (27%) |

| **Variable** | **N = 525<sup>1</sup>** |
| --- | --- |
| *Hindu* | 380 (72%) |
| *Muslim* | 1 (0.2%) |
| *Not Mentioned* | 1 (0.2%) |
| **Caste** |  |
| *General* | 19 (3.6%) |
| *Not mentioned* | 93 (18%) |
| *Other Backward Castes* | 46 (8.8%) |
| *Scheduled Caste* | 349 (66%) |
| *Scheduled Tribe* | 18 (3.4%) |
| **Education** |  |
| *No formal education* | 69 (13%) |
| *Primary School (Grades 1-5)* | 97 (18%) |
| *Middle School (Grades 6-8)* | 90 (17%) |
| *Secondary School (Grades 9-10)* | 129 (25%) |
| *Higher Secondary School (Grades 11-12)* | 62 (12%) |
| *Diploma/Vocational training* | 20 (3.8%) |
| *Graduate Degree* | 43 (8.2%) |
| *Post Graduate Degree and Above* | 15 (2.9%) |
| **Marital Status** |  |
| *Currently Married* | 424 (81%) |
| *Divorced* | 1 (0.2%) |
| *Never Married* | 36 (6.9%) |
| *Not Mentioned* | 3 (0.6%) |
| *Separated* | 2 (0.4%) |
| *Widowed* | 59 (11%) |
| **Socio-economic Status** |  |
| *Lower (V)* | 45 (8.7%) |
| *Upper Lower (IV)* | 292 (57%) |
| *Lower Middle (III)* | 129 (25%) |
| *Upper Middle (II)* | 49 (9.5%) |
| **Migration Status** |  |
| *Intra State Migration* | 105 (20%) |
| *Inter State Migration* | 7 (1.3%) |

<sup>1</sup>n (%); Mean (SD)

We used the syndromic surveillance checklist to identify the incidence of various infectious syndromes in the study sample. Of the 525 samples surveyed 194 had infection syndromes in the past 3 months. The incidence of any infection in the past 3 months was 36.95% (95%CI 32.81 – 41.24%). The distribution of various infection syndromes in the sample is shown in Figure 1. Acute respiratory infection (29.1%) was the most common, followed by Influenza Like Illness (8.4%).

**Figure 1:**
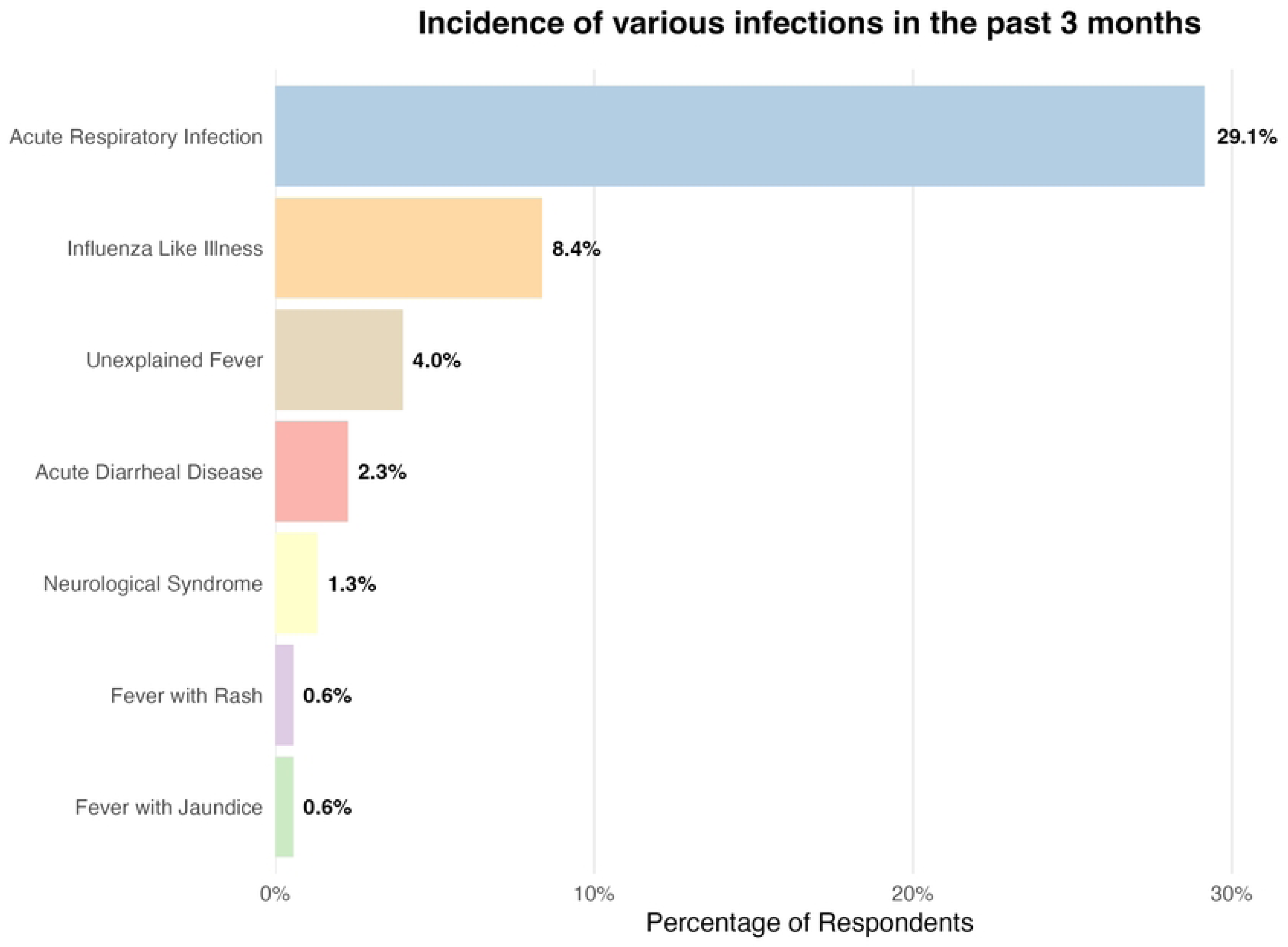
This figure shows the distribution of various infection syndromes in the study sample in the past 3 months.

Of the 194 who had infections, 143 (73.7%) sought treatment in a health facility. The distribution of the place of treatment is shown in Table 2. Of all those who sought treatment 54 (39.4%) went to a private general practitioner and 39 (28.5%) went to a government primary health center. Four persons got their medicines over the counter from a local pharmacy. The factors influencing the place of treatment is shown in Table 3. It is seen that people belonging to middle class socioeconomic status have a 3.69 (95% CI 1.60, 9.36) times greater odds of attending a private health facility compared to a government health facility after adjusting for gender, caste and migration status.

**Table 2.** Treatment Seeking Behavior for Individuals with Infection Burden.

| Treatment Facility | Frequency (N = 143)<sup>1</sup> |
| --- | --- |
| **Place of Treatment Seeking** |  |
| Government CHC | 4 (2.9%) |
| Government District Hospital | 6 (4.4%) |
| Government Medical College | 5 (3.6%) |
| Government PHC | 39 (28.5%) |
| Pharmacist / OTC | 4 (2.9%) |
| Private Clinic GP | 54 (39.4%) |
| Private Clinic Specialist | 8 (5.8%) |
| Private Hospital | 16 (11.7%) |
| Traditional Healer | 1 (0.7%) |
| Unknown | 6 |

<sup>1</sup>n (%)

**Table 3.**
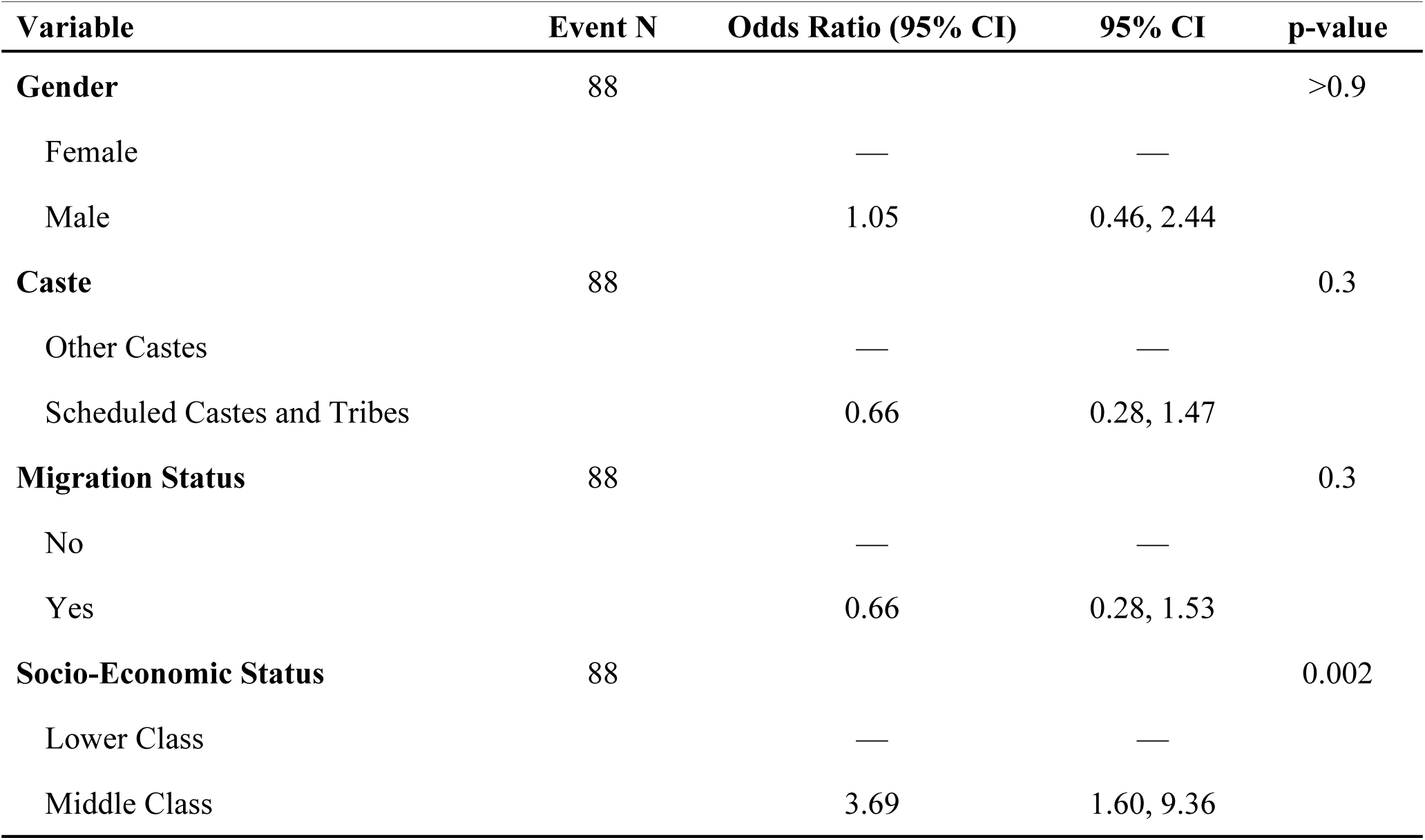
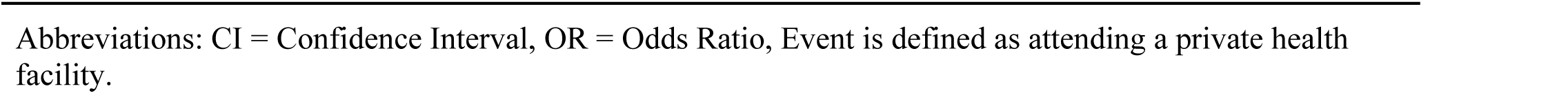
Factors Associated with Choice of Treatment Facility (Private versus Government)

| Variable | Event N | Odds Ratio (95% CI) | 95% CI | p-value |
| --- | --- | --- | --- | --- |
| **Gender** | 88 |  |  | >0.9 |
| Female |  | — | — |  |
| Male |  | 1.05 | 0.46, 2.44 |  |
| **Caste** | 88 |  |  | 0.3 |
| Other Castes |  | — | — |  |
| Scheduled Castes and Tribes |  | 0.66 | 0.28, 1.47 |  |
| **Migration Status** | 88 |  |  | 0.3 |
| No |  | — | — |  |
| Yes |  | 0.66 | 0.28, 1.53 |  |
| **Socio-Economic Status** | 88 |  |  | 0.002 |
| Lower Class |  | — | — |  |
| Middle Class |  | 3.69 | 1.60, 9.36 |  |

Abbreviations: CI = Confidence Interval, OR = Odds Ratio, Event is defined as attending a private health facility.

Of the 143 persons with infection syndromes who attended a health facility / pharmacy, 28 (19.6%) used antimicrobials. Amoxycillin (15/28), Azithromycin (6/28), Amikacin + Ceftriaxone (2/28), Piperacillin + Tazobactam (1/28), Metronidazole (1/28), Ceftriaxone (1/28), Cefotaxime (1/28) and Amoxycillin + Clavulanic Acid (1/28) were the antibiotics that were prescribed. Figure 2 shows the distribution of antimicrobials that were prescribed based on AWaRe classification of the WHO.

**Figure 2:**
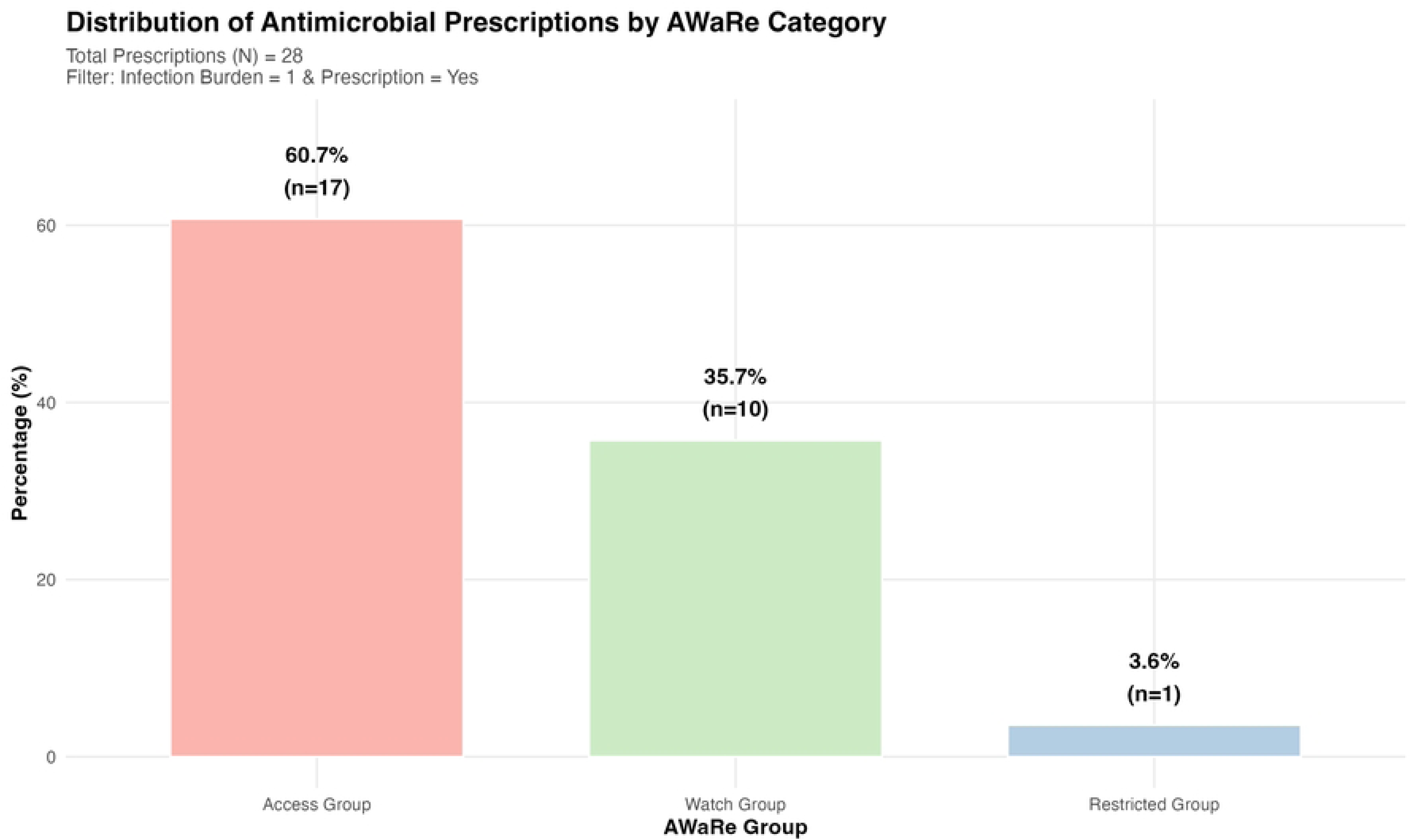
This figure shows the distribution of the antimicrobials used as per the WHO AWaRe classification.

It is seen that 60% of the prescribed antimicrobials belonged to the Access group, 35.7% to the Watch group and 3.6% to the Restrict group. Figure 3 shows the heatmap of various factors that influence receiving an antimicrobial prescription. It is seen that middle class-males- SC/ST- attending private facilities have the highest percentage of antimicrobial prescription. There are higher antimicrobial prescription densities among women and in the private sector (dark green shades in the graph). There is a high rate of antimicrobial prescription among those who did not migrate. Therefore, socioeconomic class, migration and place of treatment seem to have high effect on antimicrobial prescription.

**Figure 3:**
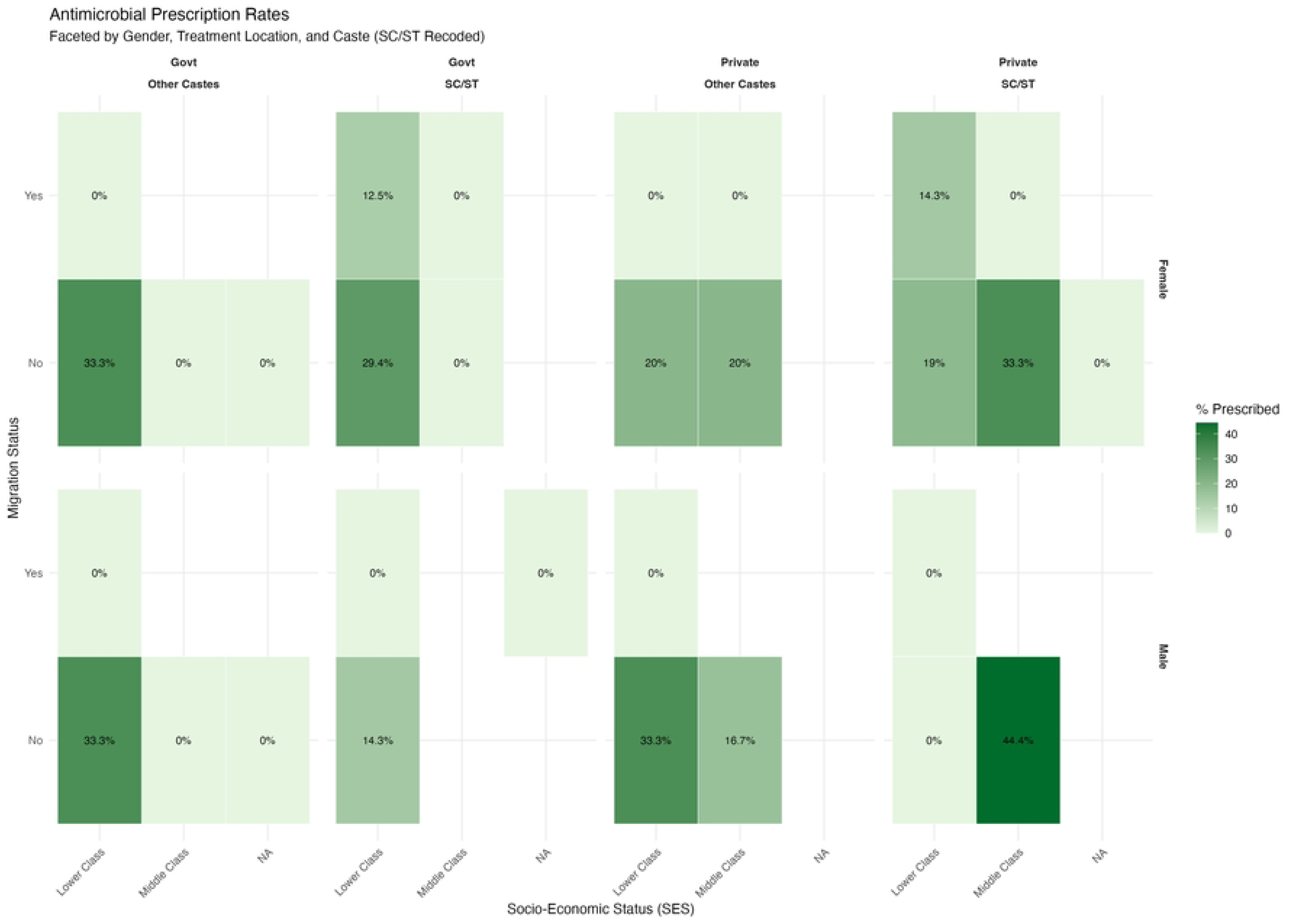
This figure shows the heatmap of antimicrobial use percentage in various shades of green across gender, socioeconomic status, caste and place of treatment.

The results of this MAIHDA analysis is shown in Table 4. The fixed-effects portion of the MAIHDA model showed an overall intercept of -1.5 (95% CI: -2.2, -0.81; p < 0.001), corresponding to an average predicted probability of 18.2% for receiving an antimicrobial prescription across all intersectional strata. The strata variance is 0.087 with a Variance Partition Coefficient of 2.58%. Only about 2.6% of the reason why people get an antimicrobial prescription is explained by their intersectional group identity. The random effects of this analysis is depicted in Figure 4. It is seen that people belonging to middle class-did not migrate-went to private facilities had the highest predicted probability (22%) of antimicrobial prescription, whereas people belonging to lower class-migrants-went to government health facilities had the lowest predicted probability (16%). Though the variance is not very high, there is a statistically significant intersectional effect of socioeconomic status, migration and place of treatment on antimicrobial prescription rates.

**Figure 4:**
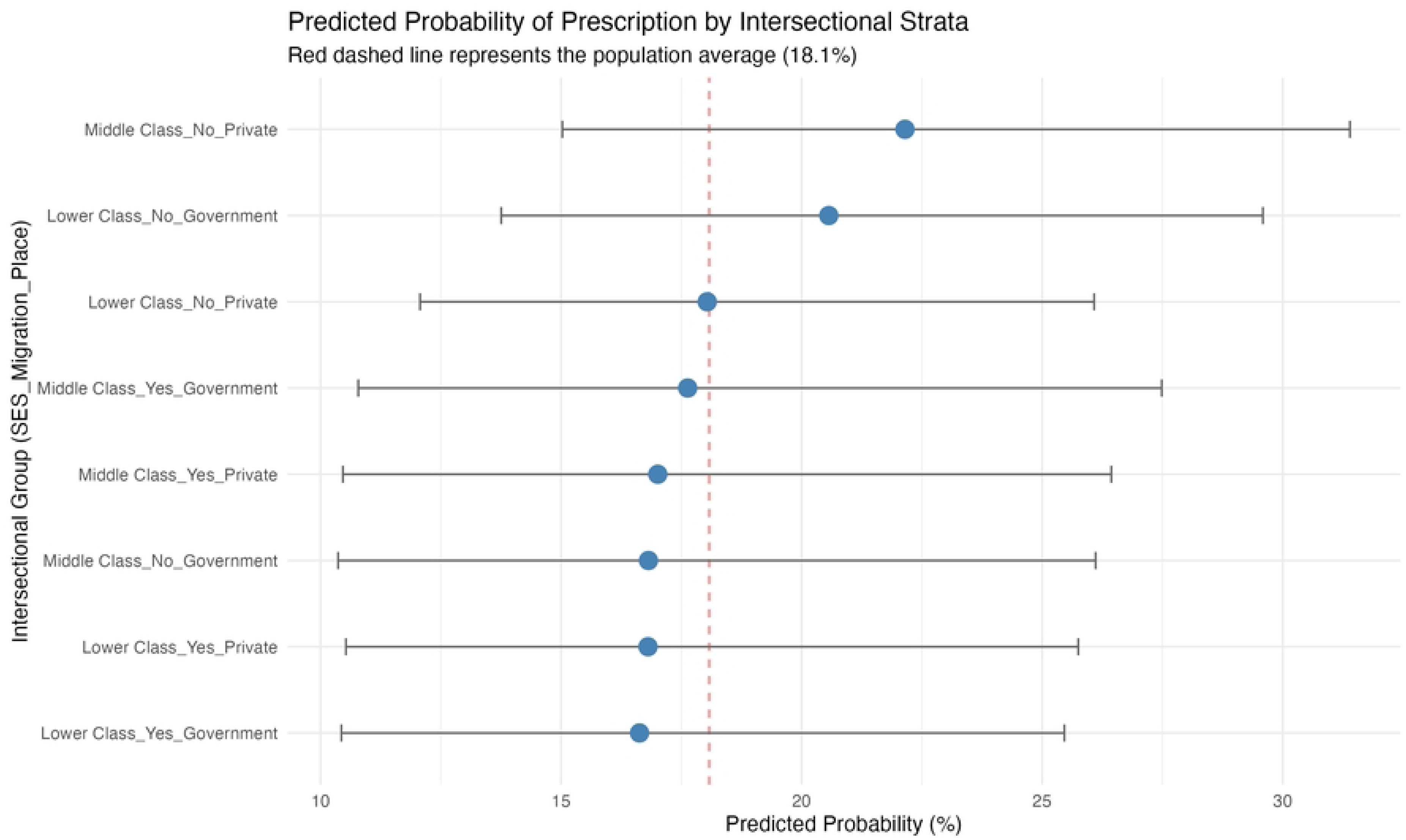
This caterpillar plot shows the predicted probability of antimicrobial use across various intersecting categories in the study sample.

**Table 4:** The intersectional effect of Socioeconomic Status, Migration and Place of treatment on antimicrobial prescription.

| Total Sample Size (N) | log(OR) | 95% CI | p-value |
| --- | --- | --- | --- |
| **Overall Population Average (Intercept)** | -1.5 | -2.2, -0.81 | <0.001 |
| No. Obs. | 139 |  |  |

Abbreviations: CI = Confidence Interval, OR = Odds Ratio
Number of Intersectional Strata: 8 | Strata Variance: 0.0871 | Variance Partition Coefficient (VPC): 2.58%

## Discussion

Most studies of antimicrobial use in India are hospital based. In this community based survey we found the incidence of infection syndromes in the past 3 months to be 37%. About 73% of those with infections sought treatment, with a majority seeking treatment in private health facilities. Those who belonged to middle class socioeconomic status had a 3.7 times greater odds of seeking treatment in private facilities compared to the lower class. Of the 143 who sought treatment for infections, 28 (19.6%) obtained a prescription / over the counter antimicrobial. Amoxycillin and Azithromycin were the most used antimicrobials. Of the antimicrobials used in the community 60% belonged to Access group 35.7% Watch and 3.6% Reserve groups as per the WHO classification. The MAIHDA model of socioeconomic status, migration and place of treatment showed a minimum but statistically significant intersectional effect on antimicrobial use with middle class – non migrant – private care seekers having significantly higher predicted probability (22%) of antimicrobial use compared to lower class – migrant – government care seekers (16%). The VPC of 2.6% indicated that the intersectional strata only explained a minor proportion of variance in the antimicrobial use.

### The following paragraphs discuss these important findings

A recent systematic review and meta analysis that included 8 studies with more than 28,000 patients showed a pooled prevalence of 65% of antimicrobial use in the primary health centre (PHC) setting.[21] A community based estimate of antimicrobial use based on pharmacy sales data showed that the rate of use of antimicrobials is 16 DID (Defined Daily Dose per 1000 persons per day), which when converted to a percentage is roughly 1.6% of the population on some antibiotic each day.[25] The most important finding from this survey is the high infection burden (37%) with a relatively low prevalence of antimicrobial use (less than 20%).

This 20% prevalence of use of antimicrobials for infections in the community setting is still high because the most common infections were acute respiratory infections and influenza like illness (37.5%) which do not require use of antimicrobials. As per the AWaRe classification by the WHO, the national level target is to have a high use of Access group greater than 60% and minimal use of the other categories of antimicrobials.[26] This study shows a high use of Access group, which is good, but an alarmingly high use of Watch antimicrobials. This is a matter for serious concern. Therefore though the issue of quantity of antimicrobial use is acceptable, the quality of antimicrobial use is problematic.

Socioeconomic class has an important effect on burden of infections and antimicrobial use. As per capita income increases people tend to move more towards private sector for their treatment, which is also seen in this study (OR 3.7). Private practitioners write prescriptions for newer and Watch group of antimicrobials, which are usually not stocked in the public sector hospitals.[21] This leads to greater use of antimicrobials determined by the socioeconomic status and place of treatment. An interesting game theory-based model showed that people belonging to lower socioeconomic class preferred to self-medicate with antimicrobials for infections. The combination of poorer access to good health care, and lack of reliable health information promotes this self-medication practice.[27] Large scale fixed effects regression analysis of time series data of antimicrobial use showed that antibiotic consumption reduced by 10.2 antibiotic doses per 1000 persons per year for every 1000 rupees increase in per capita GDP. Moreover increase in per capital government health expenditure led to reduction in private sector antibiotic use. This establishes a significant role of socioeconomic development on antimicrobial use.[28]

Analysis of a large-scale medical audit dataset of private practitioners in India, it was found that the private sector has a very high antimicrobial prescription rate of 412 per 1000 per year and a majority of them were for upper respiratory infections at the primary care level. The highest antibiotic prescription rates were observed for children below 5 years.[29] Analysis of the Kerala Medical Services Corporation data showed that the use of injectable antibiotics was more in the private sector compared to the public sector leading to use of more Watch and Reserve antibiotics. [30]

Migration leads to a situation of poor access to good quality health care services. Access to antimicrobials has a ‘dual burden’ problem.[31] People with greater access, i.e., those belonging to higher socioeconomic class, seeking treatment in private sector, have over-use of antimicrobials and those with poorer access, i.e., those belonging to lower socioeconomic class, those seeking treatment in public facilities and migrants have poor access to antimicrobials.

Besides these fixed effects of socioeconomic status, migration and place of seeking treatment, there seems to be a role for intersectional identities of individuals. This study clearly demonstrated the differences in antimicrobial use across these intersectional strata. The low Variance Partitioning Coefficient (2.6%) is common in social epidemiology. Such low VPCs indicate the presence of modest but statistically significant influence of intersectional identities. It is observed that there is a 6% (22% vs 16%) difference in probability of antimicrobial use between the two extreme strata. Such small difference can bring about huge impact at the population level. Therefore, this is not a finding that can be ignored based on its small magnitude. While 2.6% of the variance is explained by these intersectional identities various other individual level factors may influence the remaining variance. These include the nature of the infection, severity of the infection, individual prescriber characteristics, and supply side factors like availability of the antimicrobial. Among the three social factors whose intersectionality is studied only socioeconomic status is an identity of an individual. Migration status and place of seeking treatment are behavioural factors and are not particularly identities of individuals. This means that the individual’s identity may itself influence the place of seeking treatment. The low VPC must be interpreted keeping this in mind.

This study has some inherent strengths. It is one of the few community based surveys of antimicrobial use from India. It has used an advanced multi level model to study intersectional effects of social factors in influencing antimicrobial use. Migration is an important determinant of health and this is one of the few studies that has explored migration in relation to antimicrobial use. There are a few important limitations. The sample is not representative as it has a majority of women, belonging to poorer social class and scheduled castes. The survey was based on self-report of infection. The infection syndrome checklist did not capture sexually transmitted infection, soft tissue infections. Moreover, we did not capture antimicrobial use without prescription if the empty blister packs were not available for audit. It is common practice to get one or two doses of antimicrobials over the counter in these areas and this was largely missed. Despite these limitations, the study has some important findings that can influence policy.

The presence of low VPC in the MAIHDA model indicates that the focus must shift from individual level determinants of antimicrobial use to systemic and provider specific factors. There is significant antimicrobial stewardship initiatives in hospital settings. There is a need to strengthen antimicrobial stewardship at the primary care and community level. There is a need to regulate antimicrobial use in the private sector. Antimicrobial stewardship can be linked to the Clinical Establishments Act. To increase the proportion of use of Access group of antimicrobials, price capping and incentivizing the use of Access antimicrobials can be followed. Though there is a finding of low antimicrobial use in the migrant population, it need not necessarily indicate a good trend. It may indicate poor access. The health system must focus on improving health care access to migrants.

## Conclusions

This study demonstrates that antimicrobial use in the community is significantly shaped by the intersection of socioeconomic status, migrant status and place of seeking care for the infection. The presence of minimal influence of the intersectional identities, necessitates stewardship policies that target private-sector prescribing behaviors alongside broader social determinants.

## Data Availability

The anonymised data, R code and analysis outputs are available in the GitHub repository https://vijaygopichandran-cmd.github.io/Antimicrobial-data-analysis/

## Acknowledgments

The authors would like to acknowledge Ms. Ambika R, staff nurse in charge of the Omayal Achi Community Health Centre and Ms. G. Axsa, tutor Omayal Achi College of Nursing, for helping in the coordination of the survey. We would like to acknowledge the students of BSc Nursing of the 7^th^ semester batch of 2022-26 and 5^th^ semester batch of 2023-27 of the Omayal Achi College of Nursing for performing diligent and thorough data collection in the field. We would also like to acknowledge Mr. Thirunavukkarasu for support in entering the data from pen and paper format into EpiCollect 5 software.

